# Suboptimal HIV status ascertainment at antenatal clinics and the impact on HIV prevalence estimates

**DOI:** 10.1101/2021.10.04.21262474

**Authors:** Fatihiyya Wangara, Janne Estill, Hillary Kipruto, Kara Wools-Kaloustian, Wendy Chege, Griffins Manguro, Olivia Keiser

## Abstract

**Introduction:** HIV prevalence estimates is a key indicator to inform the coverage and effectiveness of HIV prevention measures. Many countries including Kenya transitioned from sentinel surveillance to the use of routine antenatal care data to estimate the burden of HIV. Countries in Sub Saharan Africa reported several challenges of this transition, including low uptake of HIV testing and sub national / site-level differences in HIV prevalence estimates.

**Methods:** We examine routine data from Kwale County, Kenya, for the period January 2015 to December 2019 and predict HIV prevalence among women attending antenatal care (ANC) at 100% HIV status ascertainment. We estimate the bias in HIV prevalence estimates as a result of imperfect uptake of HIV testing and make recommendations to improve the utility of ANC routine data for HIV surveillance. We used a generalized estimating equation with binomial distribution to model the observed HIV prevalence as explained by HIV status ascertainment and region (Sub County). We then used marginal standardization to predict the HIV prevalence at 100% HIV status ascertainment.

**Results:** HIV testing at ANC was at 91.3%, slightly above the global target of 90%. If there was 100% HIV status ascertainment at ANC, the HIV prevalence would be 2.7% (95% CI 2.3-3.2). This was 0.3% lower than the observed prevalence. Similar trends were observed with yearly predictions except for 2018 where the HIV prevalence was underestimated with an absolute bias of -0.2%. This implies missed opportunities for identifying new HIV infections in the year 2018.

**Conclusions:** Imperfect HIV status ascertainment at ANC overestimates HIV prevalence among women attending ANC in Kwale County. However, the use of ANC routine data may underestimate the true population prevalence. There is need to address both community level and health facility level barriers to the uptake of ANC services.

**Key questions:** *What is already known?:*

▪ HIV surveillance estimates from antenatal clinics (ANC) can serve as a useful proxy for HIV prevalence trends in the general female population.
▪ Kenya has conducted multiple studies which have shown that national HIV prevalence estimates from sentinel surveillance and those from routine program data to be similar.
▪ However, these studies have also revealed ongoing challenges to the suitability of using routine data as compared to sentinel surveillance including sub optimal uptake of HIV testing and sub national/ site-level differences in HIV prevalence estimates.

*What are the new findings?:*

▪ HIV positive pregnant women are more likely to be tested at ANC as compared to HIV negative women, leading to higher HIV prevalence estimates among women attending ANC.
▪ Health facility level HIV prevalence estimates are lower than that of the general population.

*What do the new findings imply?:*

▪ HIV positive women are underrepresented in antenatal clinics.
▪ In Kwale County (and similar contexts), use of routine ANC data is still not a reliable method to estimate HIV prevalence, both at facility and community level.

## Introduction

HIV prevalence estimates is a key indicator to inform the coverage and effectiveness of HIV prevention measures. For many years, countries have used sentinel surveillance data from prevention of mother to child transmission (PMTCT) or antenatal care (ANC) clinics to estimate the burden and trends of HIV infection.[1] While sentinel surveillance can provide valuable information on the burden of HIV, it is not considered representative of the general population hence may over or underestimate the general population prevalence.[2, 3] The Joint United Nations Program on HIV/AIDS (UNAIDS) and the World Health Organization (WHO) proposed transitioning from ANC sentinel surveillance to the use of routine data.[1] The latter has the advantage of expanding the representativeness of the sample while reducing logistical and financial challenges of sentinel surveillance.[4] Additionally, it is fully nested within routine health services thus women can receive a comprehensive package of care including psychosocial support and necessary referrals.[1, 5]

Kenya transitioned to the use of ANC routine test (ANC-RT) data for surveillance and aims to eliminate mother-to-child transmission (eMTCT) of HIV by 2021.[6] To achieve this, targets of 90% (ANC) attendance, 90% HIV testing among pregnant women, and 90% antiretroviral therapy (ART) use among HIV-positive pregnant women were set to be reached by 2019. The policy prescribes that HIV counseling and testing should be offered to all women at first ANC visit, with the exception of women with a previous HIV positive result.[7] The use of ANC-RT is however not without challenges and continuous data quality assessment is essential.[8, 9] Additionally, there isn’t much control on the testing and data collection procedures[10] and bias in HIV status ascertainment and subsequently prevalence may result as women can freely opt out of testing.[11]

We examine ANC-RT data for Kwale County from January 2015 to December 2019 to estimate HIV prevalence among women attending ANC at 100% HIV status ascertainment. We further estimate the absolute bias in HIV prevalence as a result of imperfect uptake of HIV testing and make recommendations to improve the utility of ANC-RT for HIV surveillance.

## Methods

This was a retrospective cross sectional study, conducted in all the four sub counties of Kwale County, Kenya.

### Data sources

We used data from the ‘Kenya Health Information System for Aggregate reporting and analysis (KHIS Aggregate)’. This is the national platform for reporting all health related data. After manual aggregation from source registers, each health facility submits monthly summary reports for validation and entry in KHIS Aggregate. We based our analysis on annual health facility-level data collected over a 5 year period; January 2015 to December 2019. The variables of interest included HIV status ascertainment and HIV positivity rates, herein referred to as HIV prevalence.

### Statistical methods

HIV status ascertainment was calculated as:

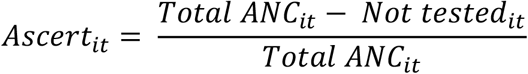

Where *Ascert*_*it*_ is the proportion of women whose HIV status was ascertained at health facility *i* in year *t*; *Total ANC*_*it*_ is the total number of pregnant women attending first ANC in that year and at the health facility; *Not tested*_*it*_ is the number of women with unknown HIV status for the same year and facility. Thus, *Ascert*_*it*_ included both newly diagnosed and previously known HIV positive women. HIV prevalence, *Pr*_*it*_, at facility *i* and year *t* was then estimated as follows:

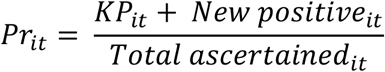

Where *KP*_*it*_ were women with a previous HIV positive result and *New positive*_*it*_ were those diagnosed as HIV positive at first ANC. Due to the longitudinal nature of the data hence likely dependence between yearly observations, we used a generalized estimating equation to model observed HIV prevalence as explained by HIV status ascertainment and region (Sub County). A binomial distribution of the form (*KP*_*it*_ + *New positive*_*it*_) ∼ *Binomial* (*Total ascertained*_*it*,_ *Pr*_*it*_) was applied. Due to the nonlinear relationship between HIV status ascertainment and HIV prevalence, the former was included in the model as restricted cubic splines with three knots. We then used marginal standardization to predict the HIV prevalence in the event the HIV status of all women attending first ANC was ascertained. Absolute bias between observed and predicted values was computed by year. Comparisons were also done across the sub counties.

Additionally, we investigated whether bias in prevalence arose due to presentation of women who were already known to be HIV positive. We fitted the same regression model as described in the preceding paragraphs, but redefined the variables as follows:

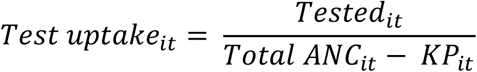

Where *Test uptake*_*it*_ is the proportion of eligible women who were tested for HIV at health facility *i* in year *t*. Subsequently, HIV prevalence among women who had no previous HIV positive result was given by

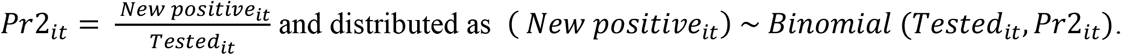

### Ethics

This study was approved by the Pwani University Ethics Review Committee.

### Patient and public involvement

It was not appropriate or possible to involve patients or the public in the design, or conduct, or reporting, or dissemination plans of our research.

## Results

A total of 139,754 pregnant women were enrolled during their first ANC visit across 124 HIV PMTCT sites over the five year period. The yearly attendance was fairly constant with a minimum of 23,509 and a maximum of 29,690 women in 2017 and 2018, respectively. The average HIV status ascertainment was at 91.3%, with 2019 and 2017 recording the least values of 85.6% and 88.7%, respectively. Observed HIV prevalence was at 3.0%, but fluctuated across the study period, from a high of 3.7% in 2017 to a low of 2.5% in 2018. HIV status ascertainment differed across the four sub counties, at 87.7%, 88.7%, 92.5% and 96.4% in Matuga, Lungalunga, Kinango and Msambweni, respectively. The observed HIV prevalence was highest in Msambweni Sub County (5.7%) and lowest in Kinango Sub County (1.6%).

Excluding known HIV positive women, 137,724 pregnant women eligible for HIV testing attended their first ANC visit within the County. The average HIV testing uptake was at 91.1%, with 2019 and 2017 recording the least values of 85.3% and 88.6%. Observed HIV prevalence was at 1.4%. Similarly, prevalence was highest in the year 2017 (2.3%) and lowest in 2018 (0.9%). Sub county trends were similar, with Msambweni recording a HIV prevalence of 3.1%. Table 1 summarizes the HIV prevalence trends across time and region.

**Table 1.**
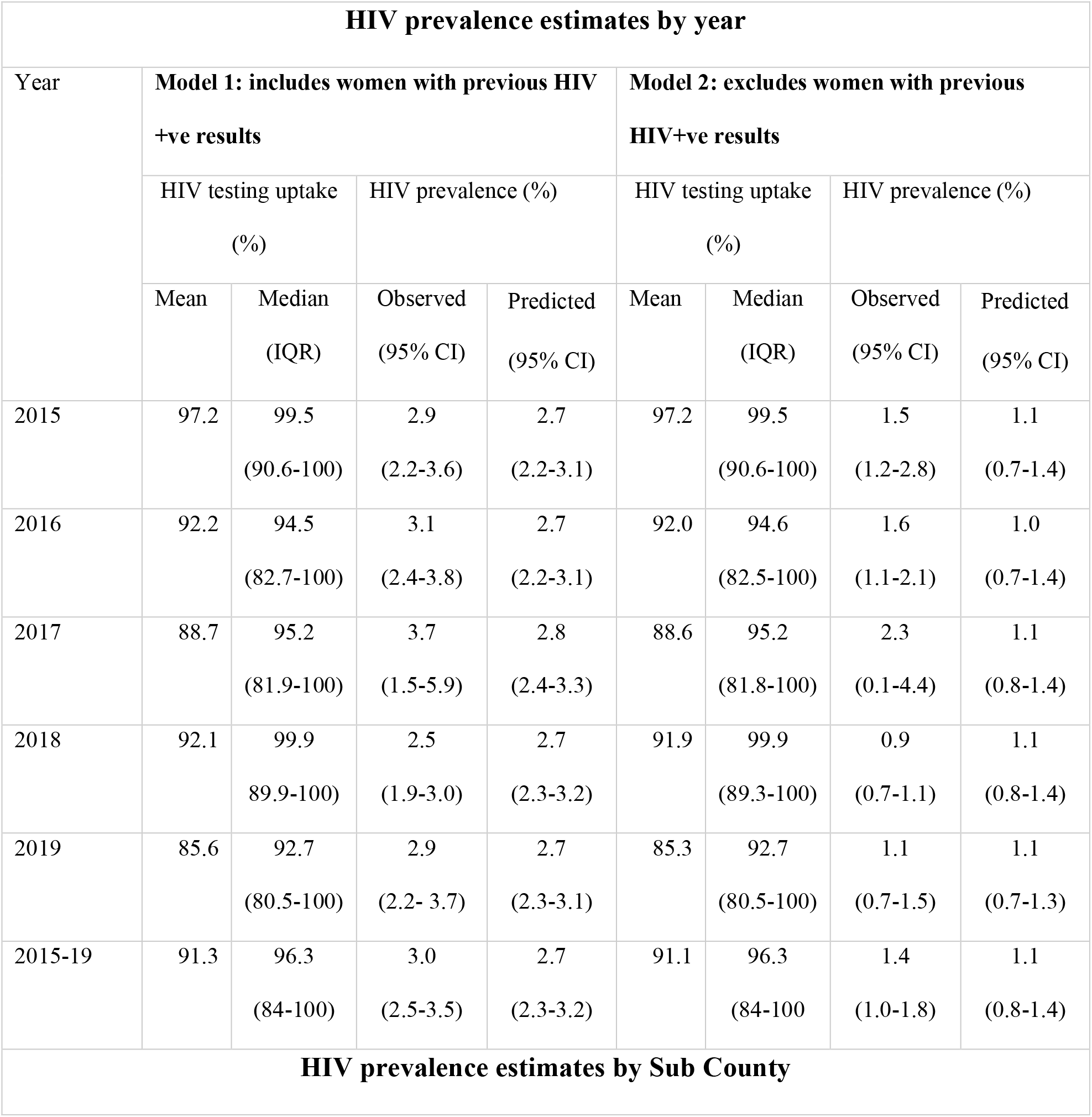
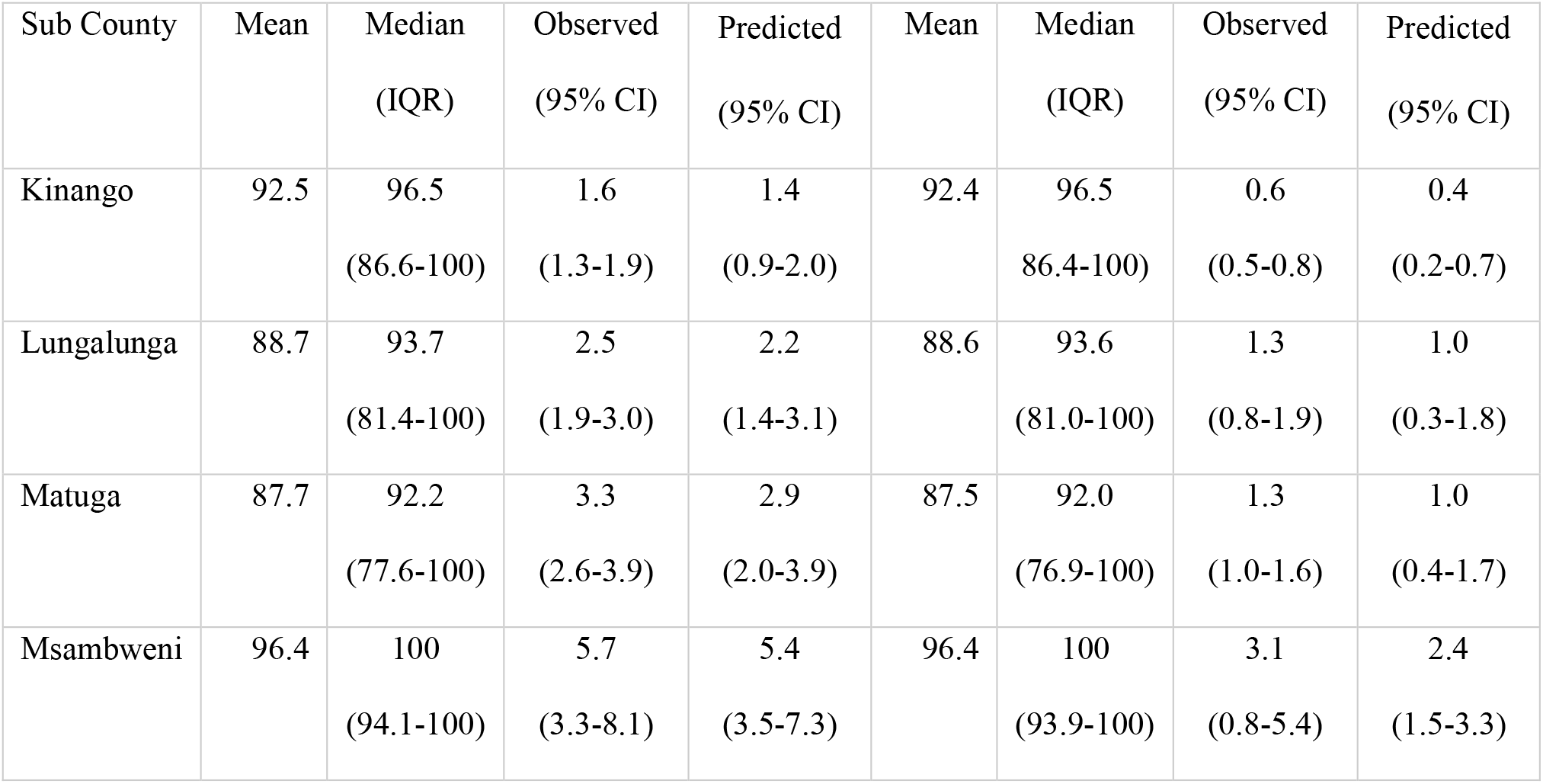
HIV prevalence estimates by year and sub county

### Predictions of HIV prevalence at 100% status ascertainment

Two models were fit; one estimating HIV prevalence among all women attending their first ANC visit and the other estimating HIV prevalence among only the women attending first ANC who were eligible for testing. The predicted prevalence was fairly constant across the study period as illustrated in (Fig.1) and (Fig.2).

**Fig. 1:**
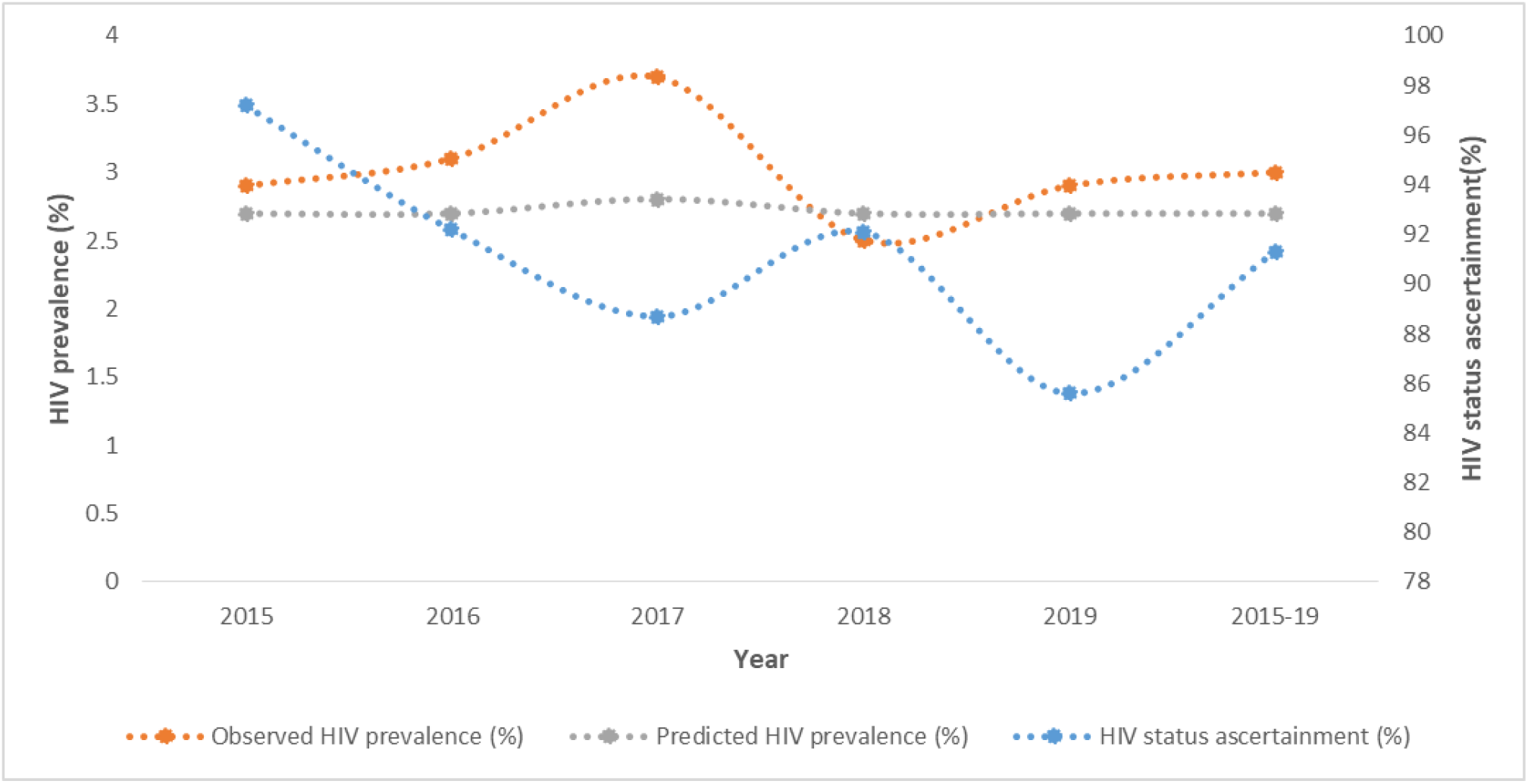
Trends of HIV status ascertainment and HIV prevalence

**Fig. 2:**
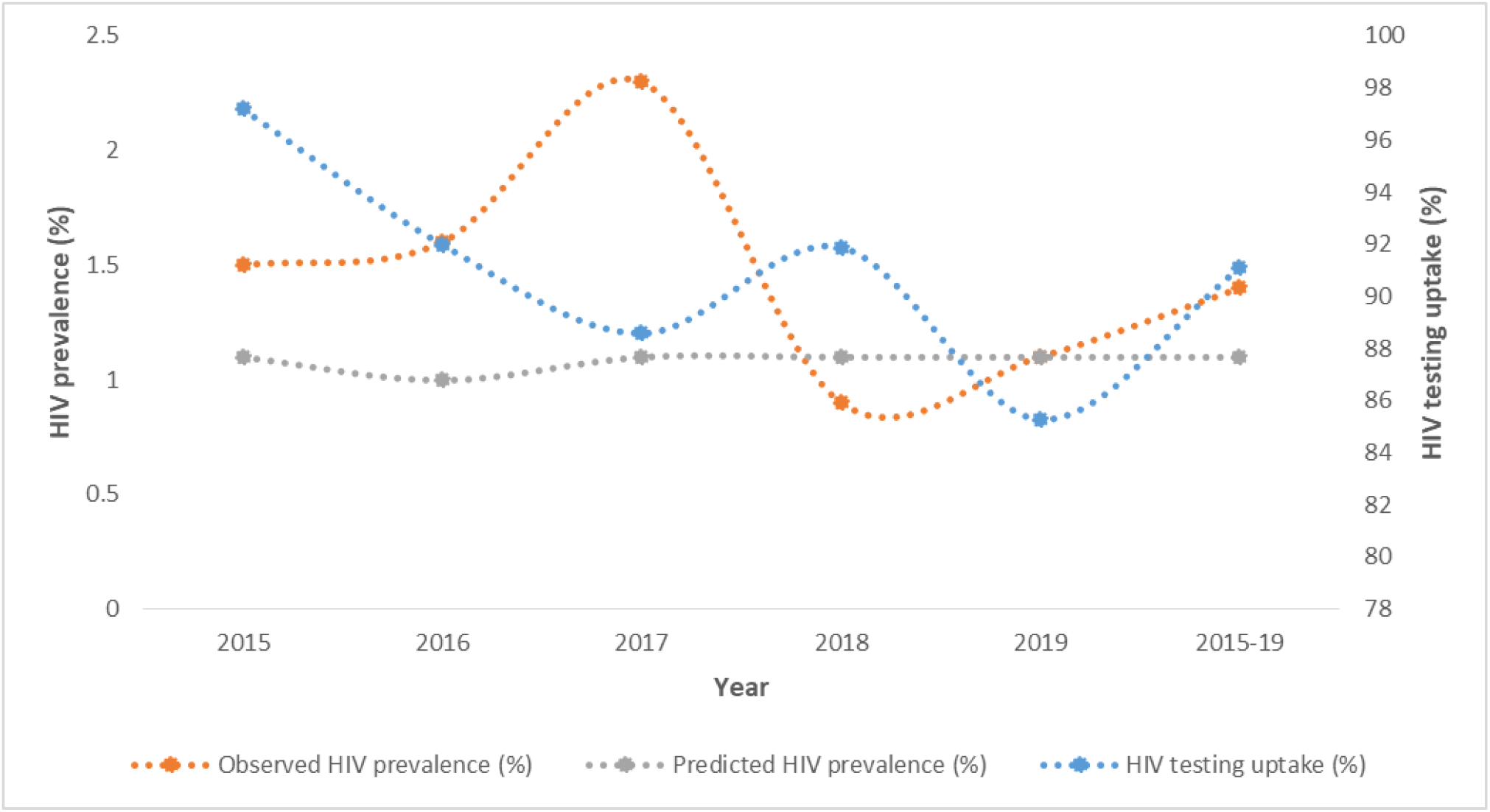
Trends of HIV testing and HIV prevalence

Uptake of HIV testing and observed prevalence did not present an obvious trend, with both measurements fluctuating over time. Notably however, the highest prevalence for both groups was recorded in 2017, alongside low uptake of HIV testing.

The model suggests that at 100% HIV status ascertainment, the prevalence would be 0.3 percent lower than the observed. Similar trends are observed with yearly predictions except for 2018 where the HIV prevalence was underestimated with an absolute bias of -0.2 percent. If there was perfect uptake of HIV test among the eligible women, the prevalence would also be 0.3 percent lower than the observed. Similarly, there were missed opportunities for identifying new HIV infections in the year 2018. This resulted in an underestimation of the prevalence by 0.2 percent. Comparison between observed and predicted prevalence at perfect testing is summarized in table 1. Additionally, both models suggest that HIV prevalence was underestimated in all the sub counties across the study period.

There was significant underestimation of prevalence in Kinango, Lungalunga and Matuga sub counties. (Fig.3) and (Fig.4) provide the means and confidence intervals of the observed and predicted prevalence. Notably, Msambweni Sub County had the highest uptake of HIV testing with comparable levels of observed and predicted HIV prevalence.

**Fig. 3:**
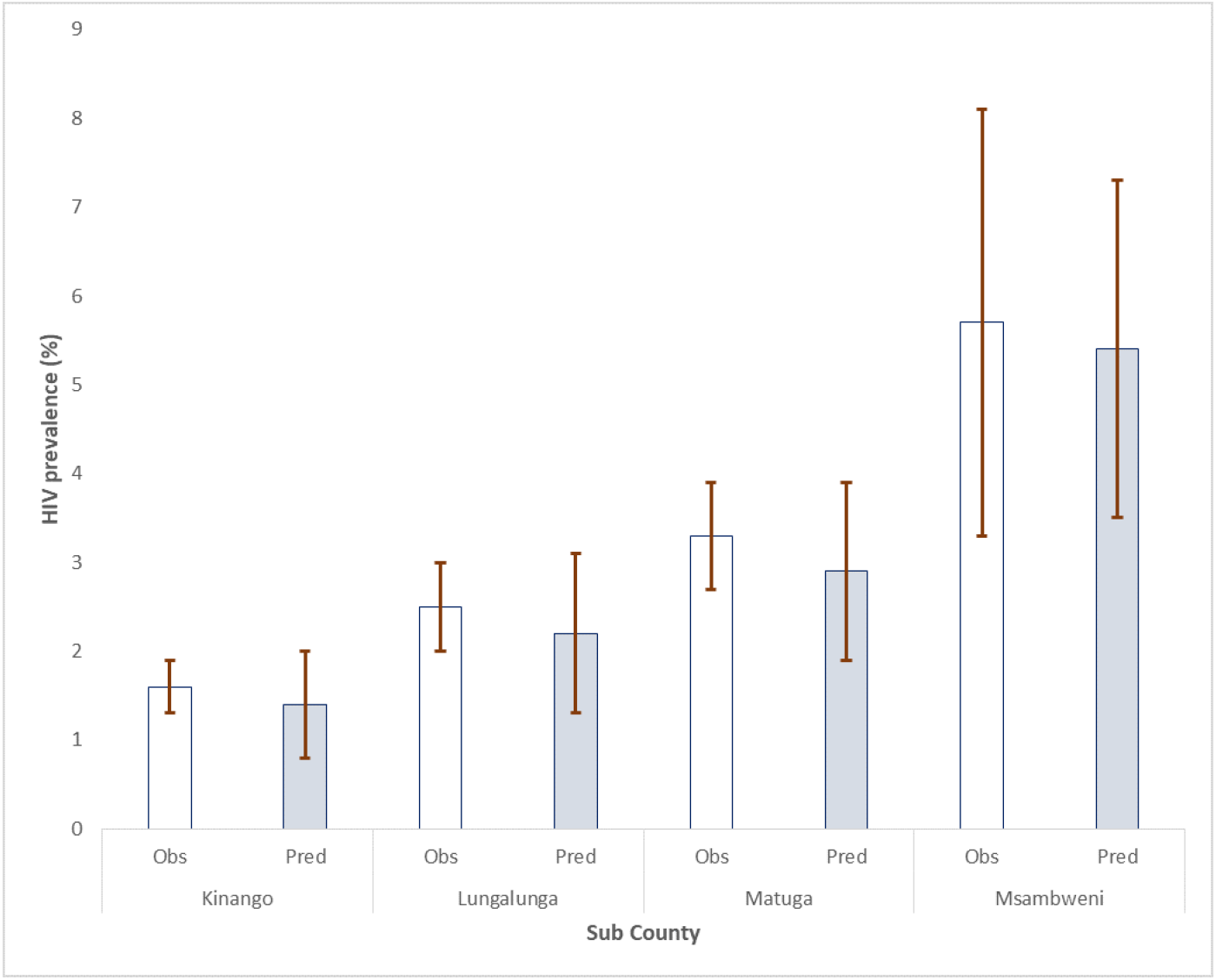
Observed and predicted HIV prevalence among all women by Sub County

**Fig. 4:**
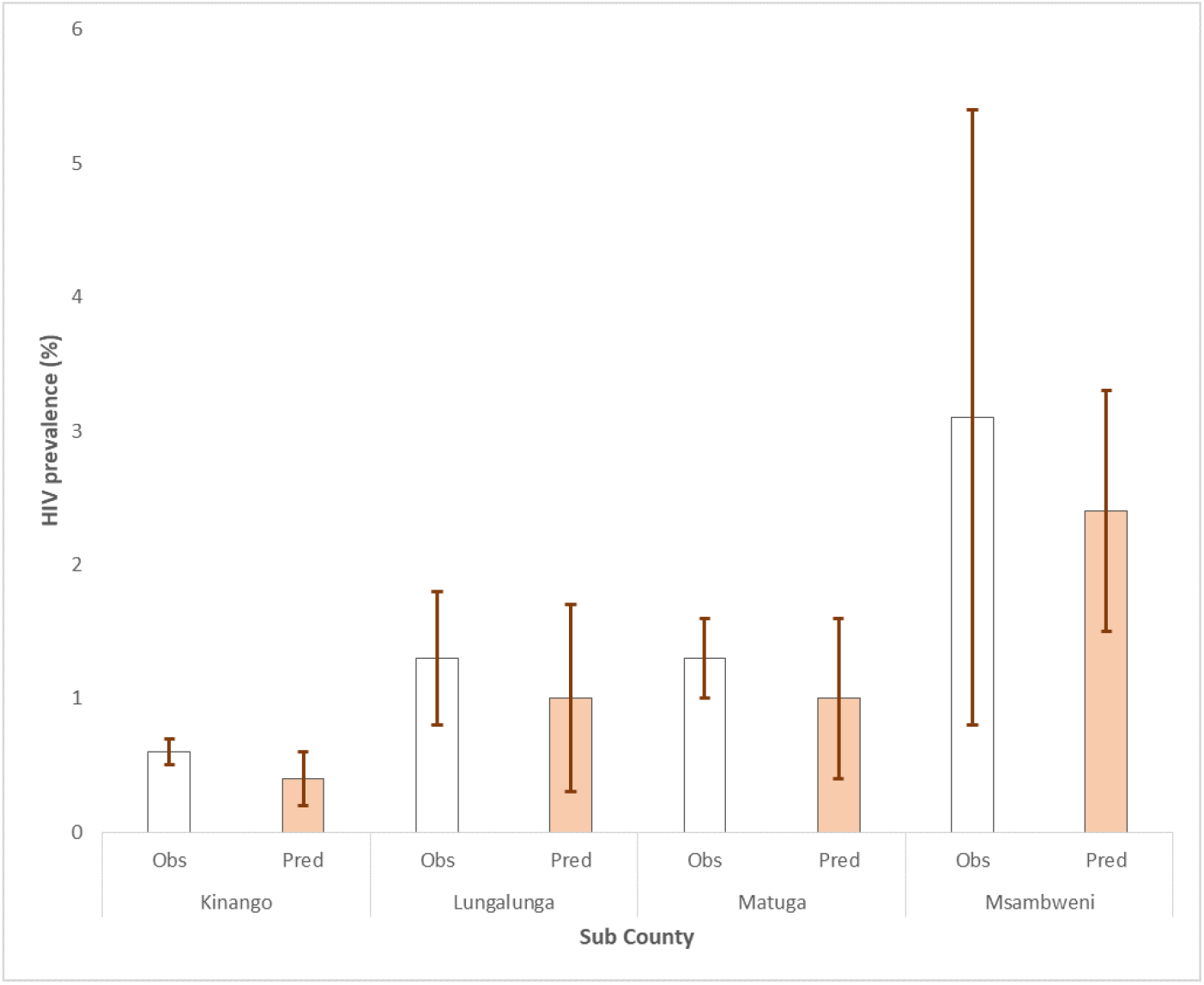
Observed and predicted HIV prevalence among newly diagnosed women by Sub County

## Discussion

The average HIV status ascertainment was at 91.3%, slightly above the UNAIDS 90% target.[12] The observed HIV prevalence was at 3.0%, but fluctuated across the study period, from a high of 3.7% in 2017 to a low of 2.5% in 2018. The year 2017 was characterized by the lowest ANC attendance, low HIV status ascertainment, and the highest observed prevalence. This suggests that women who attended ANC within that year may have been more at risk of HIV or the already HIV positive women were overrepresented. Additionally, this overrepresentation may be more profound if testing services are disrupted[11, 13] as was the case in the 2017 protracted national industrial action by healthcare workers. Excluding known HIV positive women, the average HIV testing uptake was at 91.1%, slightly lower than if all women attending ANC were considered.

The 2018 Kenya Population-based HIV Impact Assessment (KENPHIA) estimates the national and Kwale County HIV prevalence at 4.9% and 4.2% respectively.[13] The national prevalence among women of reproductive age (15 to 49 years) is estimated at 6.2% (95% CI 5.7-6.8%). While ANC data is useful in monitoring the HIV trends, it may not be representative of the general population. The lower HIV positivity in ANC suggests that HIV positive women are underrepresented at the clinics.

HIV exhibits geographical heterogeneity[14] thus contextual factors are key in understanding HIV burden estimates. In Kenya, HIV prevalence is higher in the rural (5.0%) as compared to urban areas (4.7%).[13] This is contrary to the findings of this study that show higher positivity in relatively urban sub counties of Msambweni and Matuga as compared to the lowest estimates in the relatively rural Kinango sub county. While this variation may be contributed to by differences in HIV status ascertainment, rural areas may be disproportionately affected by barriers of access to healthcare like illiteracy and socio cultural practices like early marriages.[14] Health facility level factors would also affect HIV status ascertainment and prevalence; Msambweni Sub County hosts the County referral hospital and has both the highest ascertainment and prevalence.

With adjustment for imperfect testing, the trend in predicted prevalence was fairly constant as was the case in a similar study in Malawi.[14] Imperfect HIV status ascertainment led to an overestimation of the HIV prevalence across all sub counties and years with the exception of 2018. This suggests that over the 5 year period, women who were not offered HIV testing or who opted out of testing were likely to be HIV negative. Similar findings when known HIV positive women are omitted from the analysis implies that the bias in prevalence was not due to overrepresentation of known HIV positive women. In 2018, there were missed opportunities for identifying newly diagnosed cases despite having the highest ANC attendance and ascertainment of over 90%. This implies that in 2018, women who were likely to be HIV positive opted out of testing or were not offered a HIV test. ANC prevalence estimates is evidently dependent on who gets ascertained hence the need for 100% ascertainment.[14]

Routine ANC data with optimal HIV testing has been shown to provide reliable data for monitoring the trends of HIV infection[15, 16]. Contrary findings have however been observed citing data quality and test accuracy issues in routine ANC. A 2016 study in Kenya recommended that additional preparation was required before routine antenatal HIV testing data could supplement sentinel surveillance.[8] Other studies have shown that there can be low positive percent agreement of ANC test results compared to surveillance data and recommended an assessment of the impact of site-level differences on surveillance models to be used.[17, 18]

Our study provides sub national level estimates (both county and Sub County) of HIV among pregnant women, which are normally masked when national estimates are computed.[17] Additionally we included all the PMTCT sites within the County which is more representative than a sentinel surveillance model. This study thus provides baseline information for subsequent monitoring of the PMTCT program at county level. The study was however not without limitations. Only women who attended ANC were included in the analysis hence these estimates may not be representative of all women of reproductive age within the county i.e. it excluded non pregnant women and pregnant women not attending ANC. In many parts of sub Saharan Africa, the HIV prevalence in women of reproductive age is higher than in men of the same age. In Kenya for instance, HIV prevalence of men aged 15-49 years is estimated at 2.7% (95% C1 2.4-3.1) as compared to 6.2% (95% CI 5.7-6.8) in women.[13] Thus, ANC HIV prevalence is not generalizable.

We conclude that routine PMTCT data can provide useful estimates of the burden of HIV and offers a feasible alternative to the ethical concerns raised with unlinked anonymous testing (UAT) model.[18] Imperfect HIV status ascertainment at ANC however overestimates the HIV prevalence among women attending ANC whereas the use of ANC routine data may underestimate the true HIV prevalence among women of reproductive age not attending ANC. There is thus need to address both community level (demand side) and health facility level (supply side) barriers to the uptake of ANC services if such estimates are to be reliable and more representative.

## Data Availability

The data used is available on https://hiskenya.org.

## Acknowledgements

We acknowledge the technical and logistical support offered by the County Government of Kwale as well as all HIV and RMNCAH programs implementing partners.

Author contributions: FW was responsible for all stages of the manuscript development i.e. conceptualization, methodology, data analysis, validation, write up and manuscript review. HK, JE, KW and OK all supervised the process, contributed to drafting the manuscript and final review. WC contributed to the data analysis and manuscript review. GM contributed to drafting the manuscript and final review.

## References

1. UNAIDS/WHO Working Group on Global HIV/AIDS and STI Surveillance. Guidelines for assessing the utility of data from prevention of mother-to-child transmission (PMTCT) programmes for HIV sentinel surveillance among pregnant women. WHO Guidelines 2013;52.

2. UNAIDS/WHO Working Group on Global HIV/AIDS and STI Surveillance. Guidelines for Second Generation HIV Surveillance : an update : know your epidemic. WHO Guidelines 2011;4.

3. Bezerra LM. Global report : UNAIDS report on the global AIDS epidemic 2010. Urban Res 2010;3:229–230.

4. Rennie S, Turner AN, Mupenda B et al. Conducting unlinked anonymous HIV surveillance in developing countries: Ethical, epidemiological, and public health concerns. PLoS Medicine 2009;6:30–34.

5. WHO. Consolidated guidelines on HIV testing services 2015. WHO Guidelines 2015;3–5.

6. Chirombo B, Alwar T, Yonga I, et al. Framework for Elimination of of Mother-To-Child Mother-To-Child Transmission of HIV and Syphilis in Kenya 2016-2021. 2016;1–2.

7. National AIDS and STI Control Program, Ministry of Health Kenya. Guidelines on Use of Antiretroviral Drugs for Treatment and Preventing HIV in Kenya. 2016;119–123.

8. Sirengo M, Rutherford GW, Otieno-Nyunya B et al. Evaluation of Kenya’s readiness to transition from sentinel surveillance to routine HIV testing for antenatal clinic-based HIV surveillance. BMC Infect Dis 2016;16:1–6.

9. Sheng B, Marsh K, Slavkovic AB, et al. Statistical models for incorporating data from routine HIV testing of pregnant women at antenatal clinics into HIV/AIDS epidemic estimates. AIDS 2017;31:87–94.

10. World Health Organization. March 2014 supplement to the 2013 consolidated guideline on Use of Antiretroviral Drugs for Treating and Preventing HIV Infection. 2014.

11. Tenthani L, Haas AD, Egger M et al. HIV Testing among Pregnant Women Who Attend Antenatal Care in Malawi. J Acquir Immune Defic Syndr 2015;69:610–614.

12. UN Joint Programme on HIV/AIDS (UNAIDS). 90-90-90 An ambitious treatment target to help end the AIDS epidemic. 2014.

13. NASCOP. KENPHIA 2018 preliminary report. 2020:9–12.

14. Thamattoor U, Thomas T, Banandur P et al. Multilevel analysis of the predictors of HIV prevalence among pregnant women enrolled in annual HIV sentinel surveillance in four states in Southern India. PLoS One 2015;10:7.

15. Kumar R, Virdi NK, Lakshmi PV et al. Utility of prevention of parent-to-child transmission (PPTCT) programme data for HIV surveillance in general population. Indian J Med Res 2010;132:256–259.

16. Manyahi J, Jullu BS, Abuya MI et al. Prevalence of HIV and syphilis infections among pregnant women attending antenatal clinics in Tanzania, 2011 Disease epidemiology - Infectious. BMC Public Health 2015;15:1.

17. Young PW, Mahomed M, Horth RZ et al. Routine data from prevention of mother-to-child transmission (PMTCT) HIV testing not yet ready for HIV surveillance in Mozambique: A retrospective analysis of matched test results. BMC Infect Dis 2013;13:1.

18. Kayibanda JF, Alary M, Bitera R et al. Use of routine data collected by the prevention of mother-to-child transmission program for HIV surveillance among pregnant women in Rwanda: Opportunities and limitations. AIDS Care - Psychological and Socio-Medical Aspects of AIDS/HIV 2011;23:1570–1577.

